# Modeling Joint Reference Regions for Omics Biomarkers in UK Biobank Proteomics

**DOI:** 10.64898/2026.09.01.26361504

**Authors:** Murih Pusparum, Olivier Thas, Gokhan Ertaylan

## Abstract

Conventional univariate reference intervals (UniRIs) are widely used to identify abnormal biomarker values, but they evaluate each biomarker independently and do not account for coordinated deviations between biomarkers. We developed and evaluated a joint reference region (JRR) framework for plasma proteomics data using the Olink proteomics dataset generated by the UK Biobank Pharma Proteomics Project, covering approximately 3,000 plasma proteins. JRRs were estimated for selected protein pairs in a healthy reference subset, while UniRIs were estimated separately for individual proteins using the nonparametric method. Both approaches were then evaluated in ICD-defined disease subsets. Biomarker discovery revealed sparse and heterogeneous disease–protein associations, with some proteins recurring across multiple phenotypes and others showing more disease-specific patterns. The added value of JRRs varied across diseases and protein pairs. Across evaluated protein pairs, 56.5% showed higher sensitivity under the JRR framework than the UniRI of the first protein, and 47.3% showed higher sensitivity than the UniRI of the second protein. At the disease level, the median proportion of protein pairs with improved JRR sensitivity was 0.57. JRRs were most informative when univariate detection was limited but a subset of diseased observations was flagged only by the joint region.

These findings suggest that JRRs provide a complementary approach to UniRIs by capturing abnormal joint biomarker configurations in high-dimensional proteomics data.

## 1 Introduction

Reference intervals (RIs) are the cornerstone of clinical laboratory medicine and are used routinely to interpret biomarker measurements in diagnostic and monitoring settings. Traditionally, RIs are defined as the range that includes the central 95% of values observed in a reference population considered healthy and are typically estimated separately for each biomarker using univariate statistical approaches. In clinical practice, these intervals provide a benchmark against which individual test results are evaluated, allowing clinicians to determine whether a measured value is within the expected physiological range or indicates potential pathology. Consequently, RIs play an important role in disease detection, risk stratification, and longitudinal patient monitoring across a wide range of clinical domains [1–3].

Despite their widespread use, conventional RI approaches or univariate RIs, which are typically defined separately for each biomarker, have important limitations. In particular, they treat biomarkers independently and therefore do not account for correlations between biological variables. However, many diseases are characterised by coordinated dysregulation across multiple biomarkers rather than by isolated changes in a single marker [4, 5]. Evaluating each biomarker independently thus provides only a partial view of the underlying biological state. Many physiological processes involve interactions across multiple molecular pathways, which are reflected in correlated alterations among several biomarkers. Univariate RIs therefore capture only isolated aspects of these processes and may overlook biologically meaningful patterns arising from their joint behaviour. Approaches that consider biomarkers simultaneously within a joint reference framework can provide a more integrated representation of the biological system, offering a richer context for interpreting biomarker measurements in clinical and biomedical research [6, 7].

The limitations of univariate RIs have become increasingly apparent with the rapid expansion of high-dimensional molecular profiling technologies. Advances in high-throughput omics platforms now enable the simultaneous measurement of a large number of molecular features within a single biological sample. In particular, recent developments in affinity-based proteomics technologies, such as the Olink Explore proteomics platform, allow the quantification of thousands of circulating proteins from small volumes of plasma [8, 9]. These technologies provide unprecedented opportunities to characterise complex biological processes and disease mechanisms. At the same time, large population-scale resources, such as the UK Biobank Pharma Proteomics Project, are generating proteomic measurements for tens of thousands of individuals, enabling systematic investigation of associations between circulating proteins and a wide range of disease phenotypes [10]. Together, these developments are accelerating the integration of proteomics into biomedical research, and creating strong momentum for its broader implementation in clinical practice.

However, the interpretation of such high-dimensional biomarker data remains challenging within the traditional framework of univariate RIs. When thousands of proteins are measured simultaneously in each individual, evaluating each biomarker independently fails to capture the complex correlation structure that arises from shared biological pathways, regulatory networks, and physiological processes. In this context, statistical frameworks capable of defining joint reference regions (JRRs) across multiple correlated biomarkers become increasingly important. Even joint consideration of two correlated proteins may provide a more informative representation of the biological system than evaluating each biomarker in isolation. Such joint frameworks can better reflect coordinated biological variation and offer a more biologically meaningful basis for interpreting multi-marker measurements.

Despite this need, methodological developments in reference interval estimation have remained largely within a univariate paradigm. Existing approaches typically focus on establishing thresholds for individual biomarkers, with standard clinical laboratory guidelines and reviews primarily addressing reference intervals for quantitative laboratory tests considered separately [3, 11]. Although multivariate reference-region concepts have been discussed in laboratory medicine, including reference regions of two or more dimensions and distribution-free multivariate tolerance regions, these approaches have not yet become routine for interpreting high-dimensional clinical and omics biomarker profiles [7, 12]. Moreover, there is currently no widely established statistical framework for estimating covariate-adjusted joint reference regions that are suitable for large-scale omics datasets and that can support translational interpretation in clinical and biomedical research. Addressing this gap is essential for enabling the effective integration of multi-marker proteomic measurements into clinical decision-making and precision medicine.

The objective of this study is to develop and evaluate a statistical framework for estimating joint reference regions (JRRs) for two correlated biomarkers in high-dimensional omics data. By extending the concept of univariate RIs to a multivariate setting, the proposed framework aims to capture coordinated variation across multiple biomarkers and provide a more informative representation of physiological and pathological states. To demonstrate its practical utility, we apply this framework to large-scale proteomic measurements generated using the Olink Explore proteomics platform in approximately 5,000 participants from the UK Biobank Pharma Proteomics Project. This dataset offers a unique opportunity to investigate the joint behaviour of circulating proteins across a broad range of disease phenotypes in a population-scale cohort.

The analytical workflow of this study comprises two main stages. First, we performed a biomarker discovery analysis to identify circulating proteins associated with the 100 most prevalent diseases recorded in the UK Biobank cohort. These disease-associated biomarkers are then used to construct joint reference regions (JRRs) for selected pairs of correlated proteins, allowing the modelling of multidimensional biomarker behaviour in health and disease. Subsequently, we compared the performance of the proposed JRR framework with traditional univariate RIs, evaluating their coverage properties, false positive rates, and sensitivity to detect disease-related deviations. Finally, we discuss the potential translational implications of joint reference regions for biomarker interpretation, highlighting how multidimensional representations of “normality” may improve the interpretation of omics measurements in both biomedical research and future clinical applications.

## 2 Materials and Methods

### 2.1 Data Sources

#### 2.1.1 UK Biobank Cohort

This study used data from the UK Biobank (UKB), a large prospective cohort of approximately 500,000 participants recruited between 2006 and 2010 across the United Kingdom [13]. At baseline, extensive demographic, lifestyle, clinical, and biological information was collected. Ongoing follow-up is performed through linkage to hospital records, death registries, and primary care databases. All participants provided their informed consent in writing, and ethical approval was granted by the North West Multi-centre Research Ethics Committee. Access to the data was obtained under UKB application number 71521.

#### 2.1.2 Quantitative proteomics data

The proteomic profiling was conducted by the UK Biobank Pharma Proteomics Project (UKB-PPP) using the Olink Explore platform (Uppsala, Sweden). The Olink Explore 3072 assay is based on proximity extension assay (PEA) technology, generating Normalized Protein Expression (NPX) values on a log2 relative scale. In total, approximately 54,000 UKB plasma samples were profiled, covering more than 3,000 protein biomarkers. Standard quality control procedures provided by Olink and UKB were applied, including plate-wise normalization, removal of proteins with more than 30% values below the limit of detection, and exclusion of samples failing technical QC.

#### 2.1.3 Clinical disease records

Disease outcomes were determined from linked electronic health records, including Hospital Episode Statistics (HES) for England, Scottish Morbidity Records, and Patient Episode Database for Wales. Diagnoses were coded using the International Classification of Diseases (ICD-9 and ICD-10) and procedures using the OPCS-4 coding system. For the current study, we identified the 100 most prevalent diseases in the UKB cohort, based on ICD-10 three-digit codes. The definition of cases was derived from the first diagnosis recorded in hospital records, while participants without such codes were considered controls or later referred to as the healthy reference subset.

#### 2.1.4 Clinical laboratory biomarkers

In addition to proteomic measurements, UKB provides a panel of conventional clinical biochemistry and haematological biomarkers, measured in approximately 480,000 participants at baseline [ref: UKB biomarker release]. These include standard assays such as glucose, C-reactive protein (CRP), lipid measures, and markers of renal and liver function. Where relevant, these biomarkers were incorporated for validation and comparison with proteomic-derived reference regions.

### 2.2 Methods

#### 2.2.1 Disease-biomarker association analysis

We first performed biomarker discovery analyses to identify proteomic markers associated with common diseases. The 100 most prevalent ICD-10-coded diseases were considered. For each disease, we compared biomarker levels between cases and controls using regression models adjusted for age, sex, and technical covariates (batch, plate). Proteins with missing values >30% were excluded. Associations were assessed using linear or logistic regression depending on outcome definition, and multiple testing problems were controlled using the Benjamini–Hochberg false discovery rate (FDR < 0.05). Significant biomarkers were ranked by effect size and p-value. The most consistent and strongly associated biomarkers were selected for subsequent modeling of joint reference regions.

#### 2.2.2 Modelling joint reference regions (JRRs)

We developed a statistical framework for estimating joint reference regions (JRRs), defined as the multidimensional analogue of conventional reference intervals. Unlike univariate reference intervals, which evaluate each biomarker separately, JRRs aim to capture the joint behaviour and dependency structure between two or more biomarkers. In this study, we focused on the bivariate setting, where the two biomarkers are first transformed into two variables, denoted by *Y*_1_ and *Y*_2_. These may represent the transformed protein values, after projection onto the first two principal components.

Formally, a joint reference region is defined as a two-dimensional region *R* such that

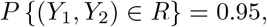

where *R* contains the central 95% of the joint distribution of (*Y*_1_, *Y*_2_) in apparently healthy individuals. The region can be written as

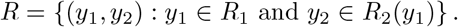

Here, *R*_1_ denotes the marginal reference interval for *Y*_1_, while *R*_2_(*y*_1_) denotes the conditional reference interval for *Y*_2_ given *Y*_1_ = *y*_1_. The marginal region for *Y*_1_ is defined by lower and upper empirical quantiles,

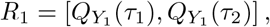

where

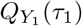

is the (100 *× T*_1_)-th percentile of the ordered values of *Y*_1_, and

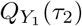

is the (100*×T*_2_)-th percentile of the ordered values of *Y*_1_. For a central 95% marginal interval, *τ*_1_ = 0.025 and *τ*_2_ = 0.975.

The conditional region for *Y*_2_ is estimated using quantile regression. Specifically, the lower and upper conditional quantiles of *Y*_2_ given *Y*_1_ are modelled as

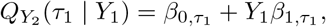

and

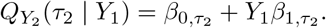

Thus, the conditional reference interval for *Y*_2_ is given by

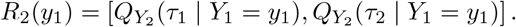

Combining the marginal interval for *Y*_1_ and the conditional interval for *Y*_2_, the estimated joint reference region is

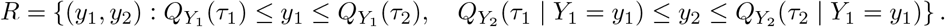

Covariates, including age, sex, and technical factors, can be incorporated into the quantile regression model to obtain covariate-adjusted JRRs. In that case, the conditional quantile model can be extended as

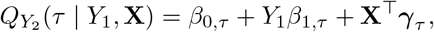

where **X** denotes the vector of covariates. Model training and evaluation were performed using a split-sample approach with internal cross-validation. The empirical coverage probability and stability of the estimated regions were assessed in apparently healthy individuals, and their performance was compared with classical univariate reference intervals. Disease subsets were then used to evaluate whether JRRs could identify abnormal joint biomarker patterns that were not captured by univariate intervals alone.

#### 2.2.3 Comparison with univariate reference intervals (UniRIs)

To evaluate the added value of the JRRs, we compared their performance with conventional univariate reference intervals (UniRIs) derived for individual biomarkers. Both methods were applied to the Olink proteomics dataset generated by the UKB-PPP, covering approximately 3,000 plasma proteins, and were estimated exclusively using individuals included in the healthy reference subset, as described in Section 2.1.3. For each pair of biomarkers, or protein pair, a JRR was constructed to characterise the expected joint distribution of the two biomarkers in apparently healthy individuals.

In parallel, UniRIs were estimated separately for each protein using the nonparametric method recommended by the International Federation of Clinical Chemistry and Laboratory Medicine (IFCC) and the National Committee for Clinical Laboratory Standards (NCCLS) for determining reference intervals, as described in [14].

The estimated JRRs and UniRIs were then subsequently evaluated in the diseased subset described in Section 2.1.3. For each observation, UniRIs classified a protein value as abnormal when it fell below the lower reference limit or above the upper reference limit. The JRRs, on the other hand, classified a protein pair as abnormal when the corresponding two-dimensional observation fell outside the estimated joint reference region. This comparison enabled us to assess whether modelling the joint distribution of two biomarkers could reveal disease-associated patterns that were not captured by evaluating each biomarker independently.

We considered three evaluation metrics. First, sensitivity was calculated as the proportion of diseased observations flagged as abnormal. For JRRs, the sensitivity was computed separately for each pair of proteins, while for UniRIs, the sensitivity was computed for each individual protein. Second, we calculated the proportion of diseased observations flagged by the JRR but not by the corresponding UniRIs, thus quantifying the additional detection attributable to the joint modelling approach. Third, we identified pairs of proteins that provided the most informative complementary interpretation relative to UniRIs. These pairs were defined as those for which the JRR highlighted joint abnormalities that were not apparent from the univariate assessment alone, supporting the potential value of JRRs for detecting coordinated biomarker deviations.

## 3 Results

### 3.1 Biomarker discovery identifies disease-associated proteins across prevalent UK Biobank phenotypes

To establish a disease-informed basis for joint reference modelling, we first performed a biomarker discovery analysis of the 100 most prevalent disease phenotypes in the UK Biobank cohort. Using Olink Explore proteomic measurements, we identified circulating proteins associated with each disease and summarised the results as a disease-by-protein presence matrix. This first stage was intended to prioritise candidate biomarkers for downstream joint modelling and to characterise the extent to which proteomic dysregulation is shared or disease-specific among common clinical phenotypes.

As shown in Figure 1, the overall pattern of detected associations was sparse. Most of the disease–protein combinations showed no selected signal, indicating that only a limited subset of the measured proteins was prioritised for any given disease. At the same time, the distribution of the detected proteins was clearly heterogeneous across phenotypes: while some diseases were associated with only a few selected proteins, others showed broader sets of associated markers. This suggests that the degree of detectable proteomic perturbation varies substantially between disease groups.

**Fig. 1.**
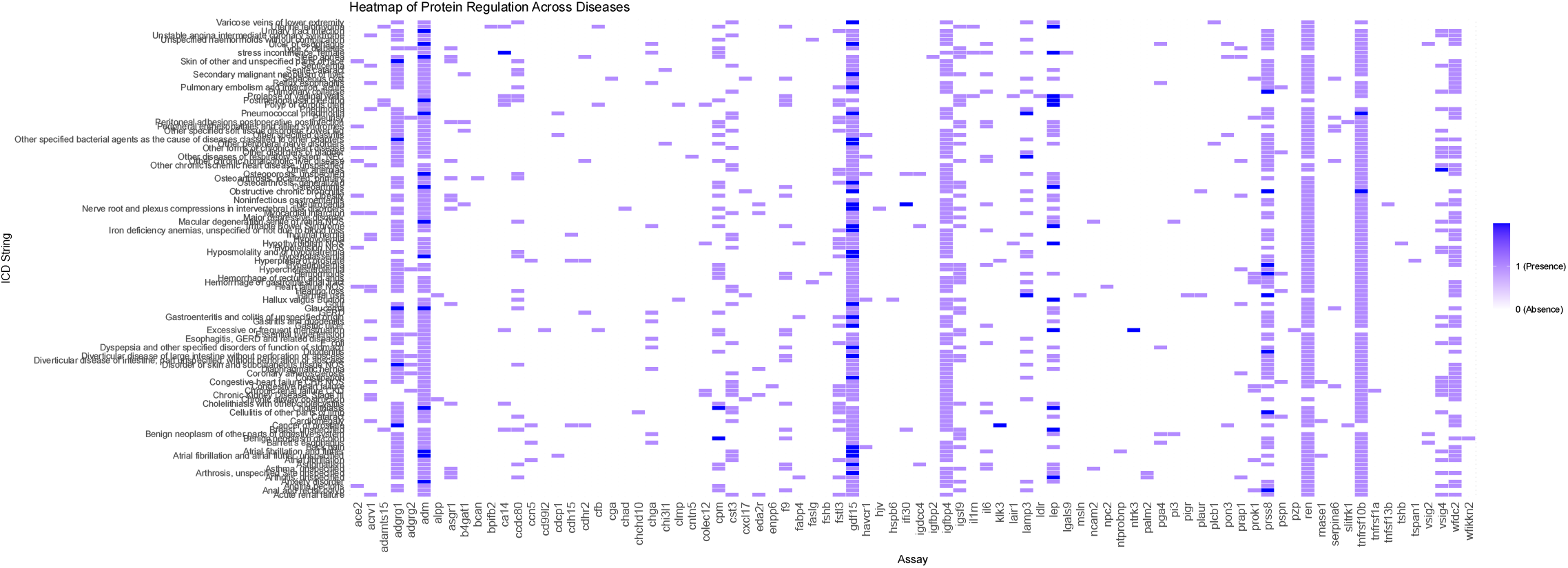
Heatmap of selected disease-protein associations across ICD groups. Rows represent diseases (ICD) and columns represent Olink plasma protein assays. Coloured cells indicate selected associations from the biomarker discovery analysis. The sparse and heterogeneous pattern suggests that detectable proteomic perturbations vary across diseases, while recurrent vertical signals indicate proteins selected across multiple phenotypes.

The heatmap also indicates that the results of the biomarker discovery were not evenly distributed among proteins. Rather than each disease being characterised by an entirely distinct set of proteins, a subset of assays appeared repeatedly across multiple phenotypes, forming vertical patterns of recurrent detection in the heatmap. This recurrence suggests that some circulating proteins may reflect shared pathological processes or general inflammatory or systemic responses. However, other proteins appeared to be more selectively associated with a smaller number of conditions.

Together, these findings support the use of a targeted pair-based strategy for downstream JRR analysis. The biomarker discovery stage identified a manageable subset of disease-relevant proteins while also revealing substantial heterogeneity in how proteomic signals are distributed across diseases. Importantly, the presence of repeatedly selected proteins and non-uniform disease-specific patterns provides a suitable basis for evaluating whether joint modelling of correlated biomarkers can capture disease-related structure beyond what is apparent from single-protein assessment alone.

### 3.2 Construction of JRRs for protein pairs

Using the healthy reference subset, we constructed JRRs for selected protein pairs identified in the biomarker discovery stage. Figure 2 shows an illustrative example of the construction procedure. Panel (a) displays the healthy reference data for one protein pair (*nefl* and *gfap*). Panel (b) shows the estimated lower and upper limits used to define the boundaries of the region: univariate limits for the first protein and conditional limits for the second protein across the observed range of the first. To obtain the final JRR, the healthy reference data were then transformed using singular value decomposition (SVD), and the region was represented in the space of the first two derived components. Panel (c) shows the resulting JRR in this transformed space, which provides the final two-dimensional representation of the healthy joint reference structure for the selected protein pair. This transformed representation was used to define the boundary of the healthy reference region for downstream classification.

**Fig. 2.**
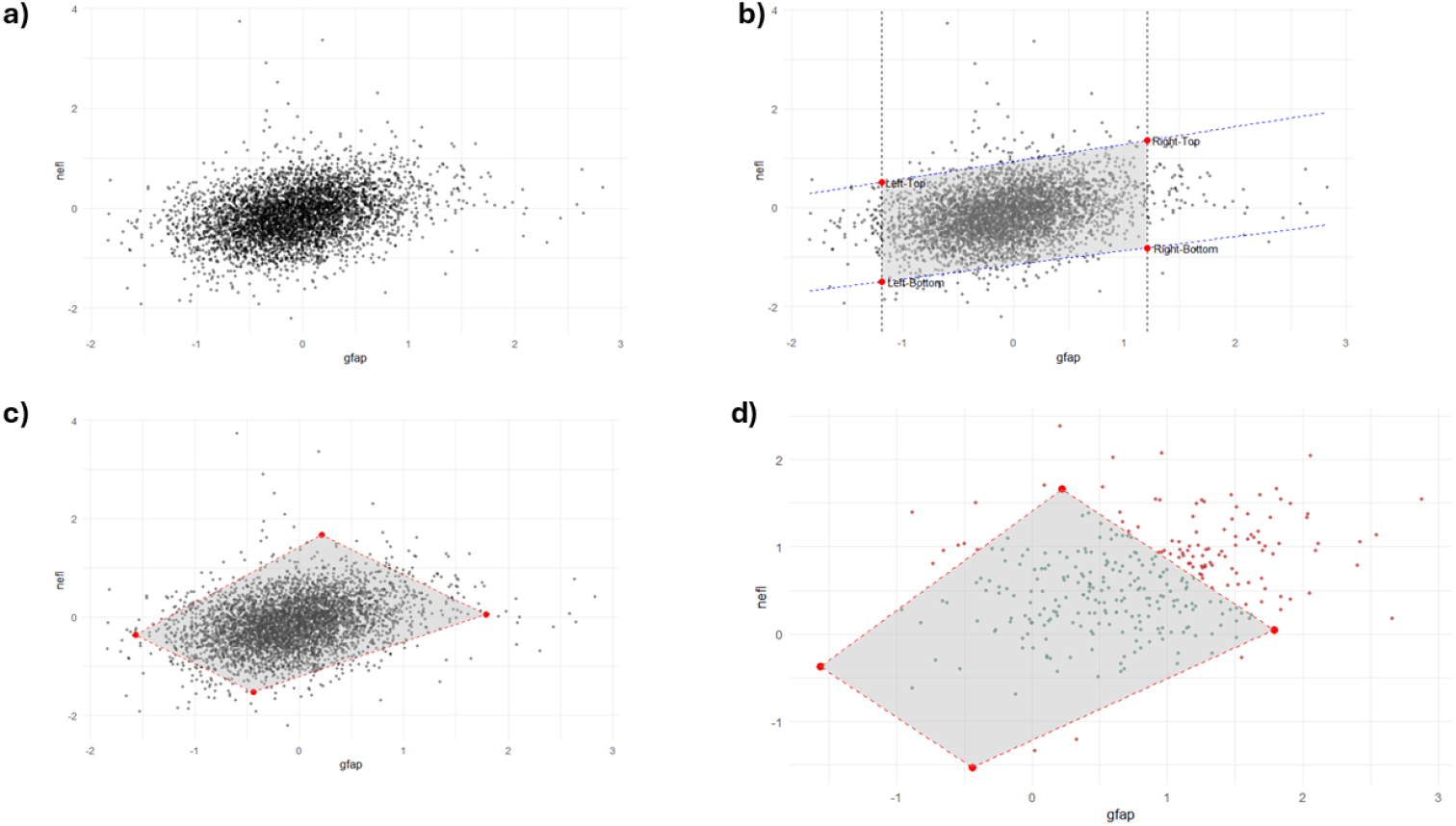
Example construction and evaluation of a joint reference region (JRR) for a selected protein pair. (a) Distribution of healthy reference observations for the example protein pair.(b) Estimation of the region boundaries based on the lower and upper limits of the first protein and the conditional lower and upper limits of the second protein. (c) Final joint reference region after transformation of the healthy reference data in the space of the first two principal components. (d) Application of the JRR to the disease subset, showing observations classified as inside or outside the healthy joint region.

The estimated JRR was subsequently applied to the disease subset to classify observations as inside or outside the healthy joint region (panel d). This evaluation step provides a direct way to assess whether disease observations show deviations in the combined protein space, and forms the basis for comparison with conventional univariate reference intervals.

### 3.3 JRRs provide complementary detection beyond UniRIs

Next, we evaluated whether JRRs improved the detection of disease-related deviations relative to conventional UniRIs. For each disease and selected protein pair, we compared the sensitivity of the JRR framework with that of the corresponding univariate approach and further examined the extent to which JRR-flagged observations overlapped or extended beyond those identified by UniRIs.

The relative sensitivity of JRRs varied between diseases and between protein pairs. As illustrated in Figure 3(a), JRR sensitivity was not uniformly higher than that of the corresponding UniRIs: for some protein pairs, the JRR yielded greater sensitivity, whereas for others the univariate approach performed similarly or better. When compared directly against the UniRI of each individual protein within the evaluated protein pairs showed higher sensitivity under the JRR framework than the UniRI of the first protein, and 47.3% showed higher sensitivity than the UniRI of the second protein (Figure 3b). Thus, the advantage of JRRs was heterogeneous rather than universal. Nevertheless, when summarised at the disease level, the median proportion of protein pairs with improved JRR sensitivity was 0.57 across ICDs (Figure 3c), indicating that in a typical disease, 57% of the evaluated protein pairs achieved higher sensitivity with JRRs than with the corresponding univariate benchmark.

**Fig. 3.**
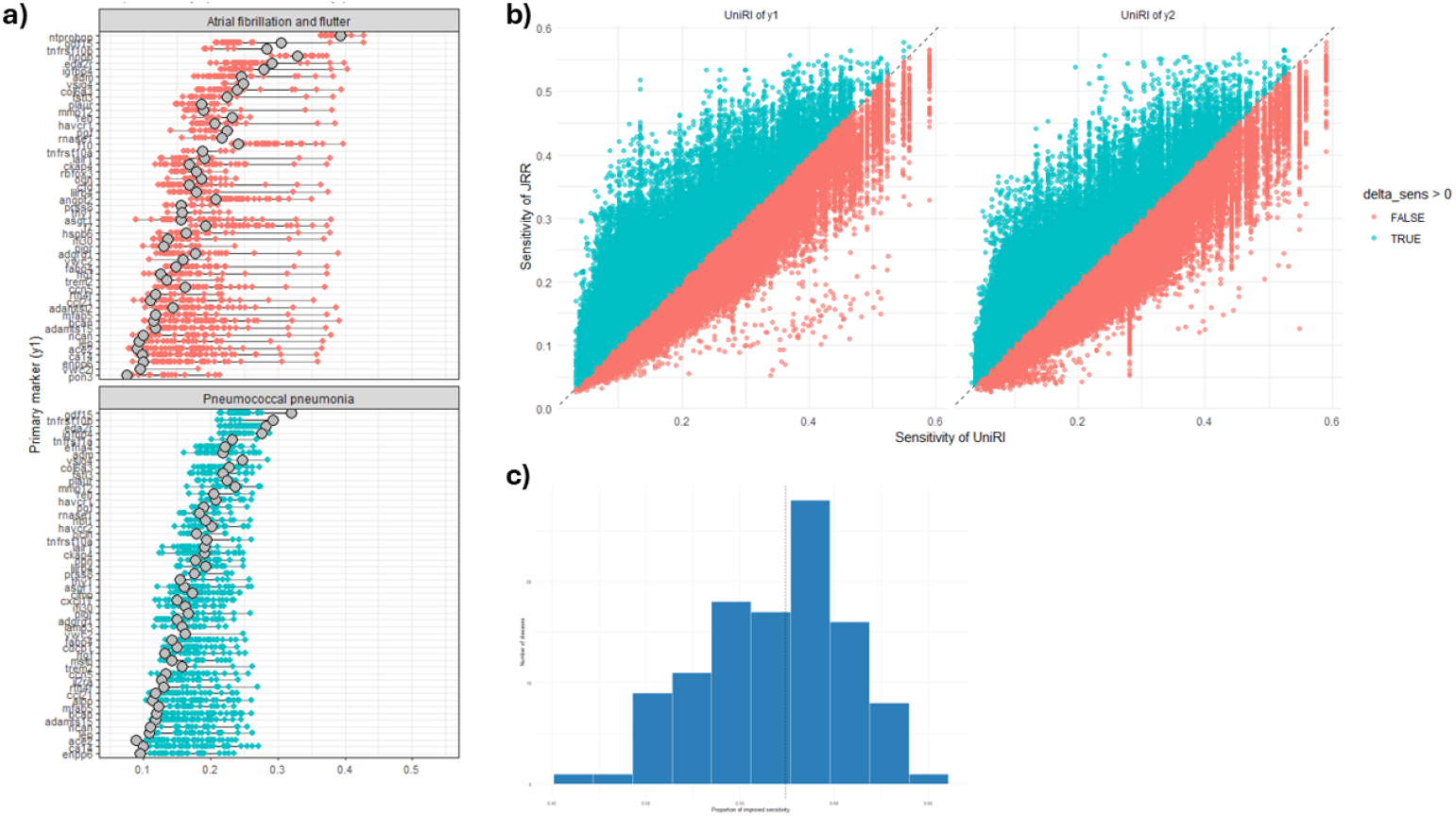
Comparison of JRR and UniRI sensitivities across diseases and protein pairs. (a) Disease-specific distribution of JRR sensitivities across evaluated protein pairs, shown relative to the corresponding UniRI sensitivities (grey points), for two example diseases. (b) Pairwise comparison of JRR sensitivity with the UniRI sensitivity of the first and second protein in each pair. Points above the diagonal indicate higher sensitivity for JRR. (c) Distribution across diseases of the proportion of protein pairs for which JRR sensitivity exceeded that of the corresponding UniRI.

Although the overall gains in sensitivity were generally modest, the JRR framework provided an important complementary perspective. Across ICDs, the proportion of cases uniquely detected by JRR was small in absolute terms, indicating that most of the abnormal observations identified by the joint framework overlapped with abnormalities already captured by at least one UniRI. However, the presence of consistent non-zero JRR-only detections across diseases shows that some disease-related deviations were detectable only when the two proteins were interpreted jointly. In other words, although the additional yield of JRRs was modest, it was not redundant.

To facilitate interpretation, ICDs were classified according to whether they showed above- or below-median values for univariate detection and unique JRR detection (Figure 4). The most favourable group for JRR complementarity consisted of ICDs with relatively low proportions flagged by UniRIs but relatively high proportions flagged only by JRR. These diseases represent settings in which marginal assessment captures comparatively little of the detectable abnormality, yet joint modelling still identifies a distinct subset of cases. By contrast, ICDs with high UniRI detection but low JRR-only detection were predominantly driven by univariate abnormalities, indicating limited additional contribution from the joint framework.

**Fig. 4.**
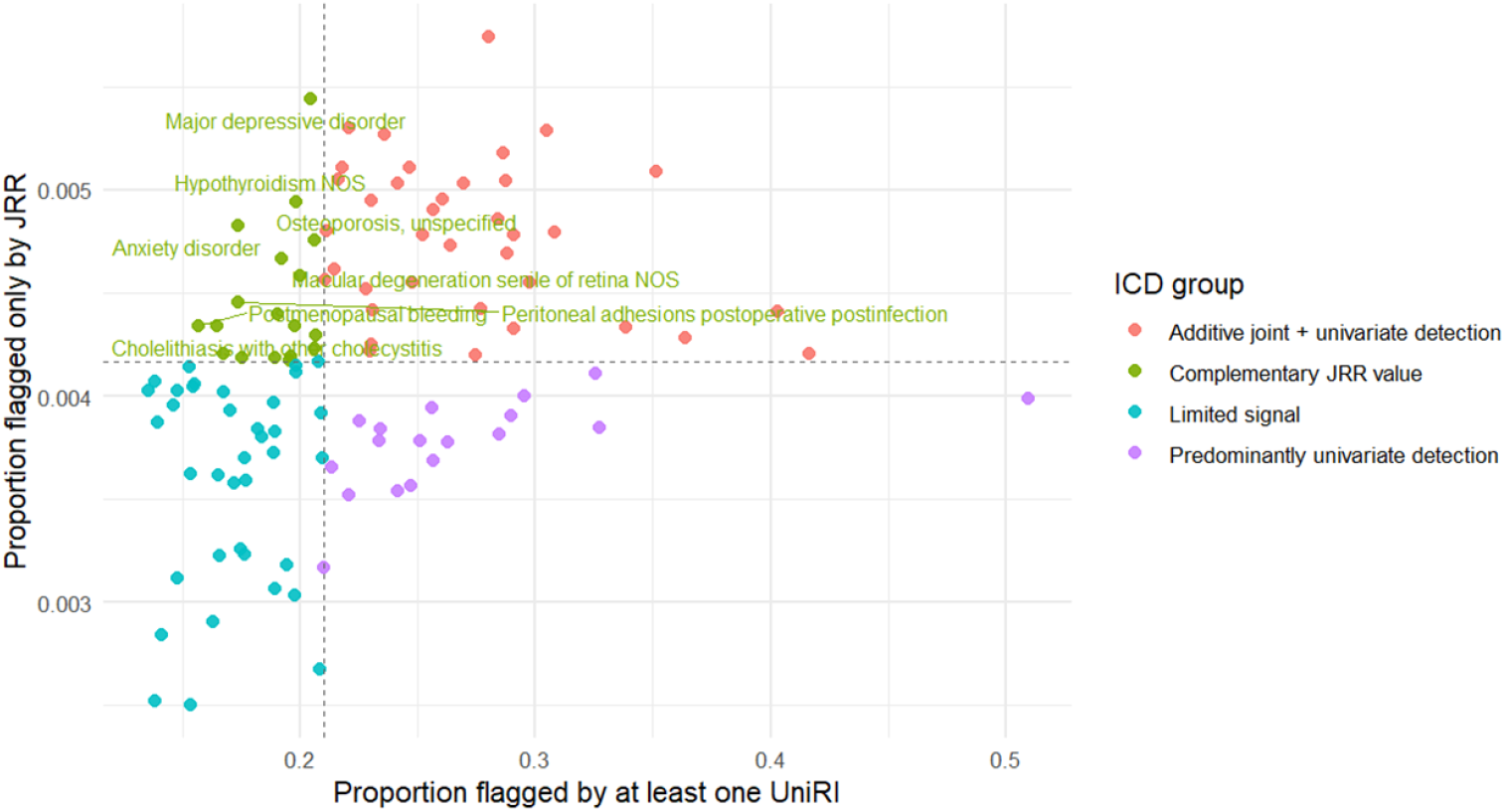
Detection patterns across ICD groups using JRRs and UniRIs. Each point represents an ICD, with the *x*-axis showing the proportion of diseased observations flagged by at least one UniRI and the *y*-axis showing the proportion flagged only by JRRs. Dashed lines indicate the median values used to classify ICD groups into four detection categories. ICD groups with low UniRI detection but high JRR-only detection were considered to show complementary JRR value, suggesting that joint biomarker modelling identifies abnormal patterns not captured by univariate assessment alone.

This pattern supports an important interpretation of the method. The main value of JRRs is not in replacing UniRIs or in producing large gains in overall detection across all diseases. Rather, JRRs appear to provide a complementary layer of interpretation by identifying combined biomarker configurations that remain within individual reference limits but are nonetheless atypical relative to the healthy joint reference space.

To further characterise the source of the complementary JRR signal, we examined pair-level detection patterns within the top ten most frequent ICD groups observed in the UKB disease subset (Figure 5). For each ICD group, protein pairs were plotted according to the proportion of diseased observations flagged by at least one UniRI and the proportion flagged only by the JRR. Across diseases, most pairs clustered near low JRR-only detection values, suggesting that the additional contribution of JRRs was limited for the majority of protein combinations. Nevertheless, selected protein pairs showed higher JRR-only detection while maintaining relatively low or moderate UniRI detection. These pairs may be considered the most informative complementary JRR candidates, as they indicate situations where the joint relationship between two proteins deviates from the healthy reference pattern even when the individual protein values are less frequently classified as abnormal by UniRIs. The labelled pairs therefore provide examples of protein combinations that may warrant closer biological interpretation or follow-up analysis.

**Fig. 5.**
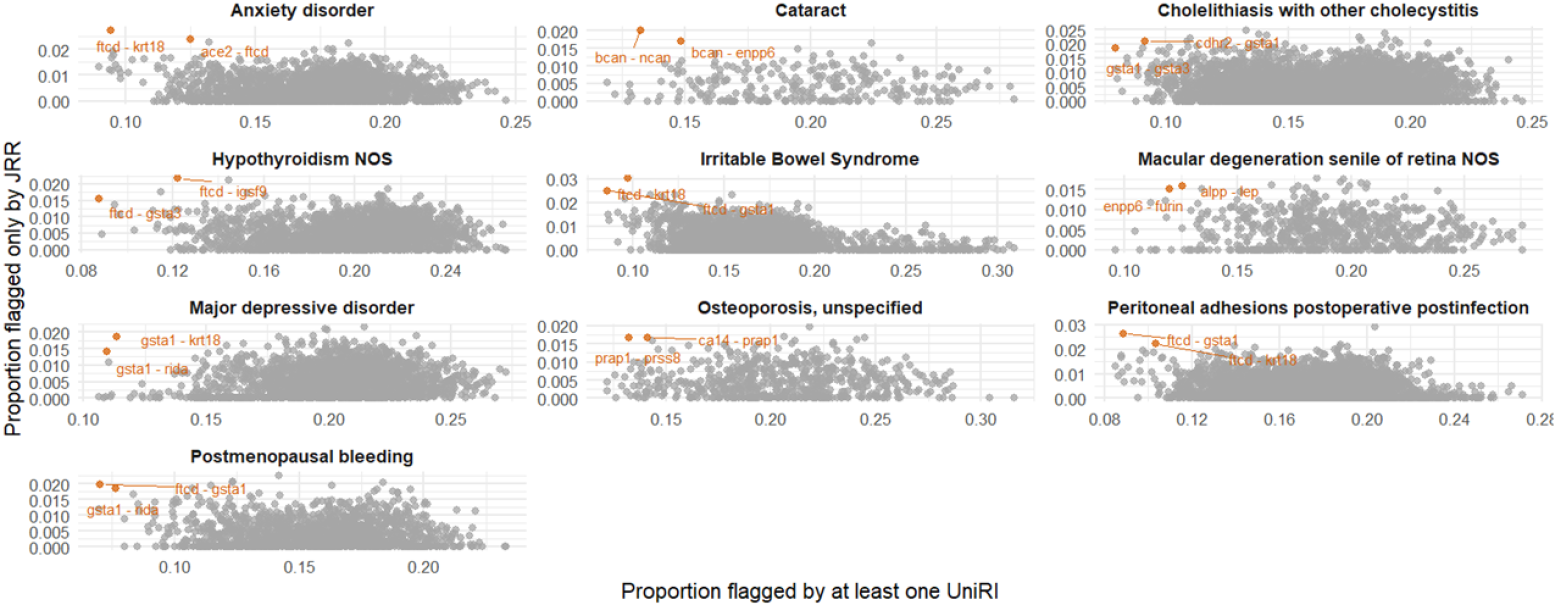
Protein pairs with complementary JRR value across the top ten most frequent ICD groups. Each panel shows pair-level UniRI detection versus JRR-only detection, with labelled points representing the most informative complementary protein pairs within each disease group.

### 3.4 Proposed framework for applying JRRs in proteomics

These results motivate a practical framework for applying JRRs in proteomics data (Figure 6). The upper panel summarises the main analytical workflow that begins with the discovery of disease-specific biomarkers to prioritise candidate proteins, followed by pair selection and construction of JRRs in a healthy reference subset. The comparative evaluation against UniRIs then enables identification of settings in which JRRs provide complementary value. In particular, the findings suggest that JRRs are most informative when disease-related abnormalities are expressed primarily in the joint biomarker configuration, while their added value is smaller when the signal is already strongly detectable by univariate assessment or when the overall pair-level signal is weak.

**Fig. 6.**
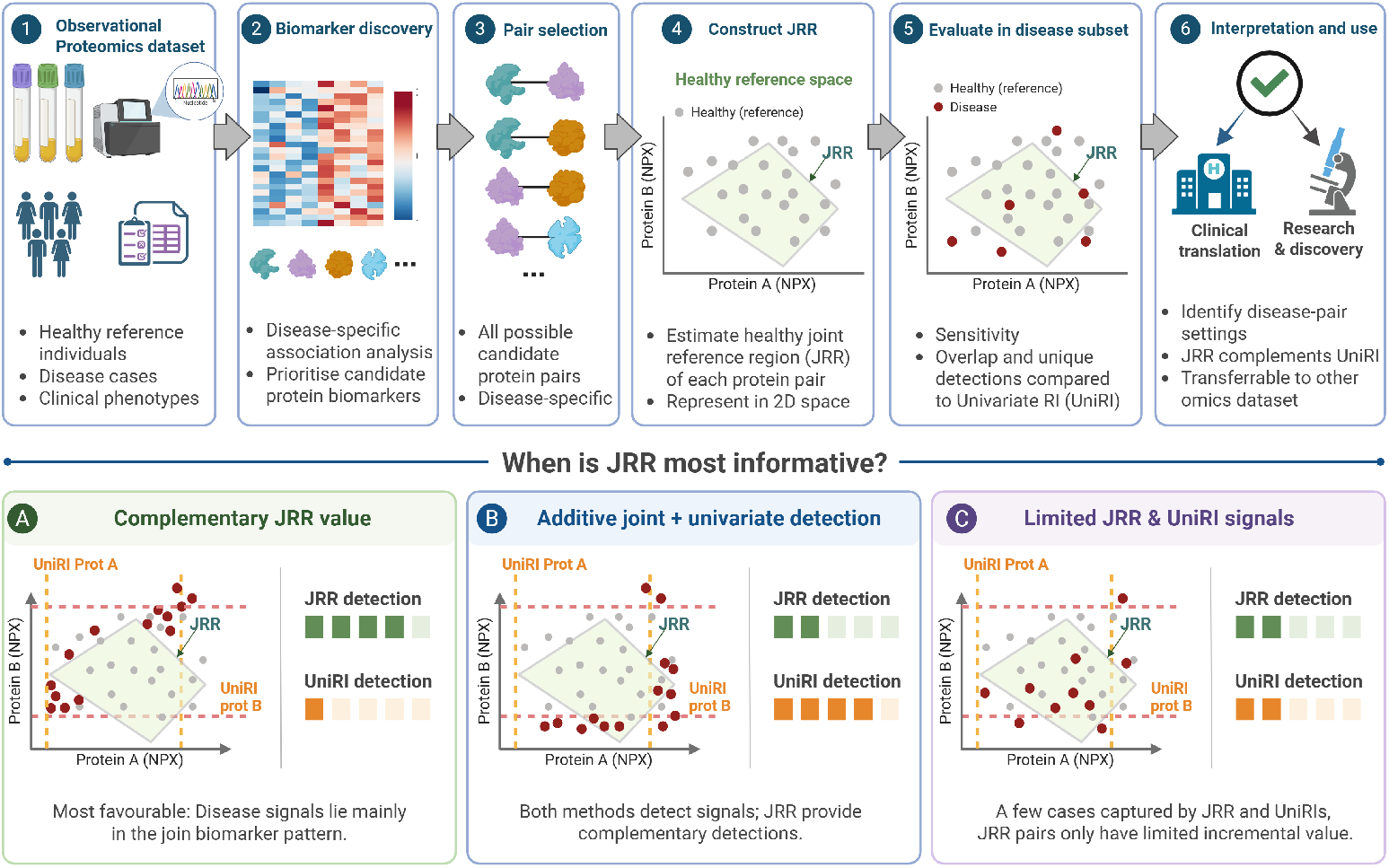
Proposed framework for applying joint reference regions (JRRs) in proteomics. The upper panel summarises the analytical workflow used in this study, including proteomics data input, disease-specific biomarker discovery, protein-pair selection, construction of JRRs in a healthy reference subset, evaluation in disease subsets, and interpretation of JRR utility relative to univariate reference intervals (UniRIs). The lower panel illustrates three main interpretive scenarios. (A) Complementary JRR value, in which UniRI detection is relatively limited but JRR adds unique detections by capturing abnormal joint biomarker configurations. (B) Additive joint and univariate detection, in which both approaches detect abnormality and JRR provides further complementary detections. (C) Limited JRR signal, in which only few additional cases are identified by JRR and the protein pair has limited incremental value.

The three lower panels of Figure 6 summarise the main interpretive scenarios observed in the analysis. Panel A represents settings with complementary JRR value, in which UniRI detection is relatively limited but JRR contributes a larger proportion of unique detections. Panel B represents additive joint and univariate detection, where both approaches detect abnormality and JRR provides additional complementary detections. Panel C represents a limited JRR signal, where only a few additional cases are identified by the joint framework. Together, these scenarios highlight that the value of JRRs depends on whether the disease-associated deviation is expressed primarily in the combined biomarker profile rather than in the marginal behaviour of individual proteins.

## 4 Discussion

In this study, we developed and evaluated a joint reference region (JRR) framework to identify disease-related deviations in high-dimensional plasma proteomics data. Using the Olink proteomics dataset generated by the UK Biobank Pharma Proteomics Project (UKB-PPP), we estimated JRRs for selected protein pairs in a healthy reference subset and compared their performance with conventional univariate reference intervals (UniRIs) in disease subsets. The main motivation was to assess whether joint modelling of protein pairs could reveal abnormal biomarker configurations that are not captured when proteins are interpreted independently. In general, our findings suggest that JRRs can provide complementary information to UniRIs, but that this added value is context-dependent and varies across diseases and protein pairs.

The comparison between JRRs and UniRIs showed that the joint framework did not uniformly outperform the univariate assessment. Instead, the relative sensitivity of the JRRs was heterogeneous: some protein pairs showed higher sensitivity using the JRRs, whereas others were captured equally or better by the UniRIs. This is expected, as JRRs are not intended to replace conventional UniRIs, but rather to identify patterns where the joint configuration of two biomarkers deviates from the expected healthy distribution. Importantly, 56.5% of the evaluated protein pairs showed higher sensitivity under the JRR framework than the UniRI of the first protein, and 47.3% showed higher sensitivity than the UniRI of the second protein. At the disease level, the median proportion of protein pairs with improved JRR sensitivity was 0.57 across ICD groups, indicating that, for a typical disease, more than half of the evaluated pairs benefited from joint modelling relative to at least one univariate benchmark. These results support the use of JRRs as a complementary layer of biomarker interpretation, particularly in settings where disease-associated alterations are expressed through coordinated protein changes rather than isolated marginal abnormalities.

A key contribution of this analysis is the distinction between different detection scenarios. Diseases with relatively low UniRI detection but higher JRR-only detection represent the clearest setting in which JRRs add value, because the univariate approach captures only a limited fraction of abnormality while joint modelling identifies additional cases. These may reflect situations which individual protein concentrations remain within their respective reference limits, but their combination is unusual relative to the healthy reference structure. In contrast, when UniRI detection is already high and JRR-only detection is low, the disease signal appears to be predominantly univariate, and the additional contribution of JRRs is limited. The pair-level analysis further showed that, although most protein pairs produced modest JRR-only detection, selected pairs within each ICD group provided more informative complementary signals. These pairs may be particularly useful for follow-up interpretation, as they highlight protein combinations in which disease-related deviation is more apparent in the joint biomarker space than in the individual biomarkers alone.

These findings also motivate a practical framework for the application of JRRs in proteomics studies. Rather than constructing JRRs exhaustively for all possible protein combinations, a more interpretable strategy is to first identify disease-associated proteins, prioritise biologically or statistically relevant pairs, and then evaluate whether their joint reference regions provide additional information beyond UniRIs. This framework is especially relevant for high-dimensional proteomics datasets, where thousands of proteins can be measured simultaneously and where disease signals may arise from coordinated biological processes. In this context, JRRs offer a way to move beyond single-marker interpretation while still retaining a reference-interval-based framework that is intuitive and clinically interpretable. The proposed classification of detection patterns, including complementary JRR value, additive joint and univariate detection, predominantly univariate detection, and limited signal, provides a useful structure for determining where joint modelling is most informative.

Several strengths of this study should be noted. First, the use of a large-scale population-based proteomics dataset allowed the estimation and evaluation of reference-based methods across a broad range of proteins and disease groups. Second, JRRs and UniRIs were estimated exclusively in the healthy reference subset and then evaluated in independent disease subsets, reducing the circularity between reference estimation and disease evaluation. Third, by comparing JRRs directly with UniRIs, the analysis provides a transparent assessment of when joint modelling adds information beyond conventional marginal thresholds. However, there are also limitations. The JRR framework was evaluated only in the bivariate setting, although many biological processes involve higher-dimensional protein networks. In addition, the observed JRR-only detection proportions were generally modest, suggesting that the incremental value of JRRs may be limited for many protein pairs and disease groups. The interpretation of protein pairs was also primarily statistical and further biological validation is needed to determine whether the identified complementary pairs reflect meaningful disease mechanisms. Finally, the disease status was defined using ICD-based groupings, which may include heterogeneous clinical phenotypes and varying disease severity.

In conclusion, JRRs provide a complementary approach for interpreting proteomics biomarkers by capturing deviations in the joint distribution of protein pairs. Although their added value is not universal, JRRs appear to be most useful in disease settings where abnormality is expressed through coordinated biomarker patterns that are not sufficiently captured by univariate reference intervals. These results support the use of JRRs as an extension, rather than a replacement, of conventional UniRIs. Future work should investigate higher-dimensional extensions, assess the robustness of JRRs across independent cohorts, and integrate biological annotation or pathway information to prioritise protein pairs with both statistical and mechanistic relevance.

## Data Availability

The individual-level UK Biobank data used in this study are not publicly available and cannot be redistributed by the authors under the terms of the UK Biobank data access agreement. Data are available to eligible researchers upon successful application to UK Biobank.

## Declarations

### Ethics approval and consent to participate

The UK Biobank has ethical approval from the North West–Haydock Research Ethics Committee as a Research Tissue Bank (REC reference: 21/NW/0157). All participants provided their informed written consent prior to participation. The present study was conducted using UK Biobank data under Application Number 71521.

### Consent for publication

Not applicable. This study used de-identified UK Biobank data and contains no identifiable participant information.

### Author contribution

Conceptualisation, M.P, O.T. and G.E.; Methodology, M.P. and O.T.; Validation, M.P. and O.T.; Software and Formal Analysis, M.P.; Writing - Original Draft, M.P., O.T., and G.E.

## Appendix A

**Section title of first appendix**

## References

[1] Horowitz, G.L.: Estimating Reference Intervals. American Journal of Clinical Pathology 133(2), 175–177 (2010) 10.1309/AJCPQ4N7BRZQVHAL. Accessed 2026-06-03

[2] Katayev, A., Balciza, C., Seccombe, D.W.: Establishing Reference Intervals for Clinical Laboratory Test Results: Is There a Better Way? American Journal of Clinical Pathology 133(2), 180–186 (2010) 10.1309/AJCPN5BMTSF1CDYP. Accessed 2026-06-03

[3] Jones, G.R.D., Haeckel, R., Loh, T.P., Sikaris, K., Streichert, T., Katayev, A., Barth, J.H., Ozarda, Y.: Indirect methods for reference interval determination – review and recommendations. Clinical Chemistry and Laboratory Medicine (CCLM) 57(1), 20–29 (2018) 10.1515/cclm-2018-0073. Accessed 2026-06-03

[4] Barabási, A.-L., Gulbahce, N., Loscalzo, J.: Network medicine: a network-based approach to human disease. Nature Reviews Genetics 12(1), 56–68 (2011) 10.1038/nrg2918. Accessed 2026-06-03

[5] Gustafsson, M., Nestor, C.E., Zhang, H., Barabási, A.-L., Baranzini, S., Brunak, S., Chung, K.F., Federoff, H.J., Gavin, A.-C., Meehan, R.R., Picotti, P., Pujana, M., Rajewsky, N., Smith, K.G., Sterk, P.J., Villoslada, P., Benson, M.: Modules, networks and systems medicine for understanding disease and aiding diagnosis. Genome Medicine 6(10), 82 (2014) 10.1186/s13073-014-0082-6. Accessed 2026-06-03

[6] Solberg, H.E.: The IFCC recommendation on estimation of reference intervals. The RefVal Program. Clinical Chemistry and Laboratory Medicine (CCLM) 42(7) (2004) 10.1515/CCLM.2004.121. Accessed 2026-06-03

[7] Boyd, J.C.: Reference regions of two or more dimensions. Clinical Chemistry and Laboratory Medicine (CCLM) 42(7) (2004) 10.1515/CCLM.2004.125. Accessed 2026-06-03

[8] Assarsson, E., Lundberg, M., Holmquist, G., Björkesten, J., Bucht Thorsen, S., Ekman, D., Eriksson, A., Rennel Dickens, E., Ohlsson, S., Edfeldt, G., Andersson, A.-C., Lindstedt, P., Stenvang, J., Gullberg, M., Fredriksson, S.: Homogenous 96Plex PEA Immunoassay Exhibiting High Sensitivity, Specificity, and Excellent Scalability. PLoS ONE 9(4), 95192 (2014) 10.1371/journal.pone.0095192. Accessed 2026-06-03

[9] Wik, L., Nordberg, N., Broberg, J., Björkesten, J., Assarsson, E., Henriksson, S., Grundberg, I., Pettersson, E., Westerberg, C., Liljeroth, E., Falck, A., Lundberg, M.: Proximity Extension Assay in Combination with Next-Generation Sequencing for High-throughput Proteome-wide Analysis. Molecular & Cellular Proteomics 20, 100168 (2021) 10.1016/j.mcpro.2021.100168. Accessed 2026-06-03

[10] Sun, B.B., Chiou, J., Traylor, M., Benner, C., Hsu, Y.-H., Richardson, T.G., Surendran, P., Mahajan, A., Robins, C., Vasquez-Grinnell, S.G., Hou, L., Kvikstad, E.M., Burren, O.S., Davitte, J., Ferber, K.L., Gillies, C.E., Hedman, K., Hu, S., Lin, T., Mikkilineni, R., Pendergrass, R.K., Pickering, C., Prins, B., Baird, D., Chen, C.-Y., Ward, L.D., Deaton, A.M., Welsh, S., Willis, C.M., Lehner, N., Arnold, M., Wörheide, M.A., Suhre, K., Kastenmüller, G., Sethi, A., Cule, M., Raj, A., Alnylam Human Genetics, AstraZeneca Genomics Initiative, Biogen Biobank Team, Bristol Myers Squibb, Genentech Human Genetics, GlaxoSmithKline Genomic Sciences, Pfizer Integrative Biology, Population Analytics of Janssen Data Sciences, Regeneron Genetics Center, Kang, H.M., Burkitt-Gray, L., Melamud, E., Black, M.H., Fauman, E.B., Howson, J.M.M., Kang, H.M., McCarthy, M.I., Nioi, P., Petrovski, S., Scott, R.A., Smith, E.N., Szalma, S., Waterworth, D.M., Mitnaul, L.J., Szustakowski, J.D., Gibson, B.W., Miller, M.R., Whelan, C.D.: Plasma proteomic associations with genetics and health in the UK Biobank. Nature 622(7982), 329–338 (2023) 10.1038/s41586-023-06592-6. Accessed 2026-06-03

[11] CLSI, Horowitz, G.L.: Defining, Establishing, and Verifying Reference Intervals in the Clinical Laboratory, Third edition edn. CLSI document EP28-A3c/C28-A3. Wayne, PA: Clinical and Laboratory Standards Institute, ??? (2010)

[12] Liu, W., Bretz, F., Cortina–Borja, M.: Distribution–free hyperrectangular tolerance regions for setting multivariate reference regions in laboratory medicine. Statistics in Medicine 43(8), 1604–1614 (2024) 10.1002/sim.10019. Accessed 2026-06-03

[13] Sudlow, C., Gallacher, J., Allen, N., Beral, V., Burton, P., Danesh, J., Downey, P., Elliott, P., Green, J., Landray, M., Liu, B., Matthews, P., Ong, G., Pell, J., Silman, A., Young, A., Sprosen, T., Peakman, T., Collins, R.: UK Biobank: An Open Access Resource for Identifying the Causes of a Wide Range of Complex Diseases of Middle and Old Age. PLOS Medicine 12(3), 1001779 (2015) 10.1371/journal.pmed.1001779. Accessed 2026-06-02

[14] Linnet, K.: Nonparametric Estimation of Reference Intervals by Simple and Bootstrap-based Procedures. Clinical Chemistry 46(6), 867–869 (2000) 10.1093/clinchem/46.6.867. Accessed 2026-06-03

